# Suggestibility in functional neurological disorder: A meta-analysis

**DOI:** 10.1101/2020.05.30.20117705

**Authors:** Lillian Wieder, Richard J. Brown, Trevor Thompson, Devin B. Terhune

**Affiliations:** Department of Psychology, Goldsmiths, University of London, London, UK; School of Health Sciences, University of Manchester, Manchester, UK; Complex Trauma and Resilience Research Unit, Greater Manchester Mental Health NHS Foundation Trust, Manchester Academic Health Sciences Centre, Manchester, UK; School of Human Sciences, University of Greenwich, London, UK

## Abstract

**Objective:** Responsiveness to direct verbal suggestions (suggestibility) has long been hypothesized to represent a predisposing factor for functional neurological disorder (FND) but previous research has yielded conflicting results. The aim of this study was to quantitatively evaluate whether FND patients display elevated suggestibility relative to controls via meta-analysis.

**Methods:** Four electronic databases were searched in November 2019, with the search updated in April 2020, for original studies assessing suggestibility using standardized behavioural scales or suggestive symptom induction protocols in FND patients and controls. The meta-analysis followed Cochrane, PRISMA, and MOOSE guidelines. Data extraction and study quality coding were performed by two independent reviewers. Standardized suggestibility scores and responsiveness to symptom induction protocols were used to calculate standardized mean differences (*SMD*s) between groups.

**Results:** Of 26,643 search results, 19 articles presenting 11 standardized suggestibility datasets (FND: *n* = 316; control: *n* = 360) and 11 symptom suggestibility datasets (FND: *n* = 1285; control: *n* = 1409) were included in random-effects meta-analyses. Meta-analyses revealed that FND patients displayed greater suggestibility than controls on standardized behavioural scales (*SMD*, 0.48 [95% CI, 0.15, 0.81]) and greater responsiveness to suggestive symptom induction (*SMD*, 1.39 [95% CI, 0.92, 1.86]). Moderation analyses presented mixed evidence regarding the extent to which effect sizes covaried with methodological differences across studies. No evidence of publication bias was found.

**Conclusions:** These results corroborate the hypothesis that FND is characterized by heightened responsiveness to verbal suggestion. Atypical suggestibility may confer risk for FND and be a cognitive marker that can inform diagnosis and treatment of this condition.

## Introduction

*Functional neurological disorder* (FND) is characterized by impaired motor or cognitive functioning that resembles neurological pathology but is not adequately explained by it.^1^ FND has a prevalence of 4–12 per 100,000^2, 3^ and represents ∼16% of clinical referrals and visits to epilepsy clinics.^4^ It is associated with considerable diagnostic delays and frequent misdiagnosis,^5^ which add to the already significant psychological, social, and economic impact of the condition.^6^

FND has long been hypothesized to be characterized by elevated responsiveness to direct verbal suggestions (suggestibility).^7^ Suggestibility is theorized to confer vulnerability for FND^8^ through aberrant meta-awareness of intentions,^9–12^ the capacity for suggestions to trigger automatized behavioural routines or mental representations,^8^ and/or a tendency to form precise (symptom) priors that override motor and perceptual systems.^13, 14^ The use of suggestion to provoke FND symptoms is widely used to aid diagnosis^15^ and functional symptoms are responsive to suggestion-based treatments, such as hypnosis and placebo.^16^ In addition, suggestibility has been shown to predict prognosis^17^ and response to treatment^18^ in FND patients. Conditions with germane symptom profiles, such as dissociative disorders, are also characterized by elevated hypnotic suggestibility.^19^

Despite these various strands of evidence, the empirical association between suggestibility and FND seems to be highly variable.^20, 21^ In order to quantify the evidence for elevated suggestibility in FND patients, we conducted a random-effects meta-analysis of controlled studies of suggestibility on standardized behavioural scales and in response to suggestive symptom induction protocols. Secondary analyses investigated moderating influences on patient-control differences, such as experimenter blindness,^22^ methodological quality, the inclusion of a hypnotic induction,^23^ and symptom provocation method.^15^

## Method

### Eligibility criteria

Inclusion criteria included (1) English language; (2) full paper in a peer-reviewed journal; (3) patient sample characterized by FND/symptoms, encompassing conversion disorder (DSM), dissociative neurological disorder (ICD), specific functional neurological syndromes (e.g., non-epileptic seizures), and conditions where functional neurological symptoms are a diagnostic feature (i.e. DSM-IV somatization disorder; Briquet’s syndrome); (4) inclusion of a control group; and either (5) use of a standardized behavioural measure of direct verbal suggestibility^23^, or (6) assessment of symptom induction through suggestion (e.g., suggestive seizure induction^15^).

Exclusion criteria included (1) studies in which suggestion was used to aid diagnosis; (2) case studies/series or non-empirical papers; (3) overlapping/insufficient data; (4) use of interrogative suggestibility scales, which capture a different form of suggestibility characterized by high compliance^24^; and (5) studies of patients with functional somatic syndromes not specifically characterized by functional neurological symptoms (e.g., fibromyalgia).

### Search strategy

PubMed, PsycINFO, Web of Science and Academic Search Complete databases were searched in November 2019 for eligible studies using terms relating to suggestibility and FND (see **online supplemental content 1**) and then integrated into a single database. The search was repeated in April 2020 but yielded no new studies. The reference lists of all eligible studies (and relevant review papers) were manually searched to identify additional studies. Authors were contacted when data were unavailable or to clarify ambiguities in methodology.

### Study selection

Two raters (LW and a second rater) independently screened and assessed all studies for their eligibility using a two-stage procedure. First, all titles and abstracts were screened and articles not meeting eligibility criteria were rejected. Second, all remaining papers were reviewed to establish a final list of articles. Discrepancies at either stage were resolved in consultation with a third reviewer (DBT) and sometimes a fourth reviewer (RJB). Authors of eligible studies were contacted to address any questions regarding insufficient data. Of 5 author groups contacted, 4 (80%) provided data sufficient to permit study inclusion or answered queries that justified exclusion.

### Data extraction

The two outcome types (standardized suggestibility and symptom suggestibility) were measured using continuous and categorical measures, respectively. After exclusion of two studies with overlapping data, data from eligible studies were extracted and coded independently by LW and the second rater using a data extraction form including: (i) study details (title, year, geographical location), (ii) diagnosis, (iii), diagnosis method, (iv) demographics (sample size and gender and age distributions), (v) study design details (suggestibility type [standardized or symptom], inclusion of a hypnotic induction, scale, administration method [live or recorded presentation], and scoring method [self or experimenter]), provocation method [see **Table 2**], and experimenter blindness [blind, unblind, or unreported]), and (vi) descriptive statistics (*M*s and *SD*s [standardized suggestibility] or response counts [symptom suggestibility]). Symptom suggestibility response counts include only responses identified as typical for the respective patient (typicality reported: *k* = 8; unreported: *k* = 3). When studies reported results for more than one provocation method (*k* = 2), the rounded mean was used. Two studies included data for both standardized and symptom suggestibility. There was 91% agreement between the two raters and discrepancies were resolved with a third reviewer (DBT).

### Study quality

A 13-item scale was developed to assess study quality (see **online supplemental content 1**). Items were adapted from an earlier meta-analysis that included items based on Cochrane criteria and PRISMA recommendations^25^ and a range of other methodological criteria such as experimenter blindness. LW and the second rater independently rated each item categorically (0 = criterion not met, 1 = criterion met) and a summed total was computed for each study. Agreement between raters was 90% (kappa = .73) and discrepancies were resolved with DBT.

### Meta-analysis and meta-regression

Individual study effect sizes included between-group differences in suggestibility that were computed with standardised mean differences (*SMD*s; Hedges’s *g*s) using Review Manager (RevMan v. 5.3, 2014; The Nordic Cochrane Centre, The Cochrane Collaboration, Copenhagen). Symptom suggestibility data consisted of binary outcomes that were used to compute odds ratios, which were subsequently transformed to *SMD*s^26^ in MATLAB (v. 2017b, MathWorks, Natick, MA). *SMD*s were coded such that positive values reflected greater suggestibility in FND patients than controls.

Publication bias was assessed by examining funnel plots of effect sizes against standard errors for asymmetry, as might occur due to a small number of studies with small or non-significant effect sizes. We also tested for asymmetry using Egger’s bias test^27^ where *p*<.05 is indicative of asymmetry. We report revised effect sizes correcting for asymmetry using the trim-and-fill method^28^ and funnel plots in **Figure 3**.

Random-effects meta-analyses were performed in JASP (v. 0.8.6, 2019; JASP Team, Netherlands). Outlier detection was made on the basis of studentized residuals (>|3.3|).^29^ There were no outliers in the standardized suggestibility data (range: −1.63 to 1.33) or the symptom suggestibility data (range: −2.01 to 1.26). Moderating effects were assessed using meta-regression analyses whenever data were available for at least 2 studies at each level of a categorical moderator and at least 10 studies for continuous moderators.^30^ Moderators included five categorical measures (experimenter blindness, hypnotic induction, symptom provocation method [suggestion vs. nocebo; symptom studies], symptom typicality [not reported vs. report; symptom studies]; and control type [non-clinical vs. clinical; standardized studies]) and one continuous measure (methodological quality).

## Results

### Study inclusion

A PRISMA diagram showing study selection is presented in **Figure 1**. Principal features of these studies, including diagnosis criteria and procedures, are presented in **Tables 1** and **2**.

**Figure 1.**
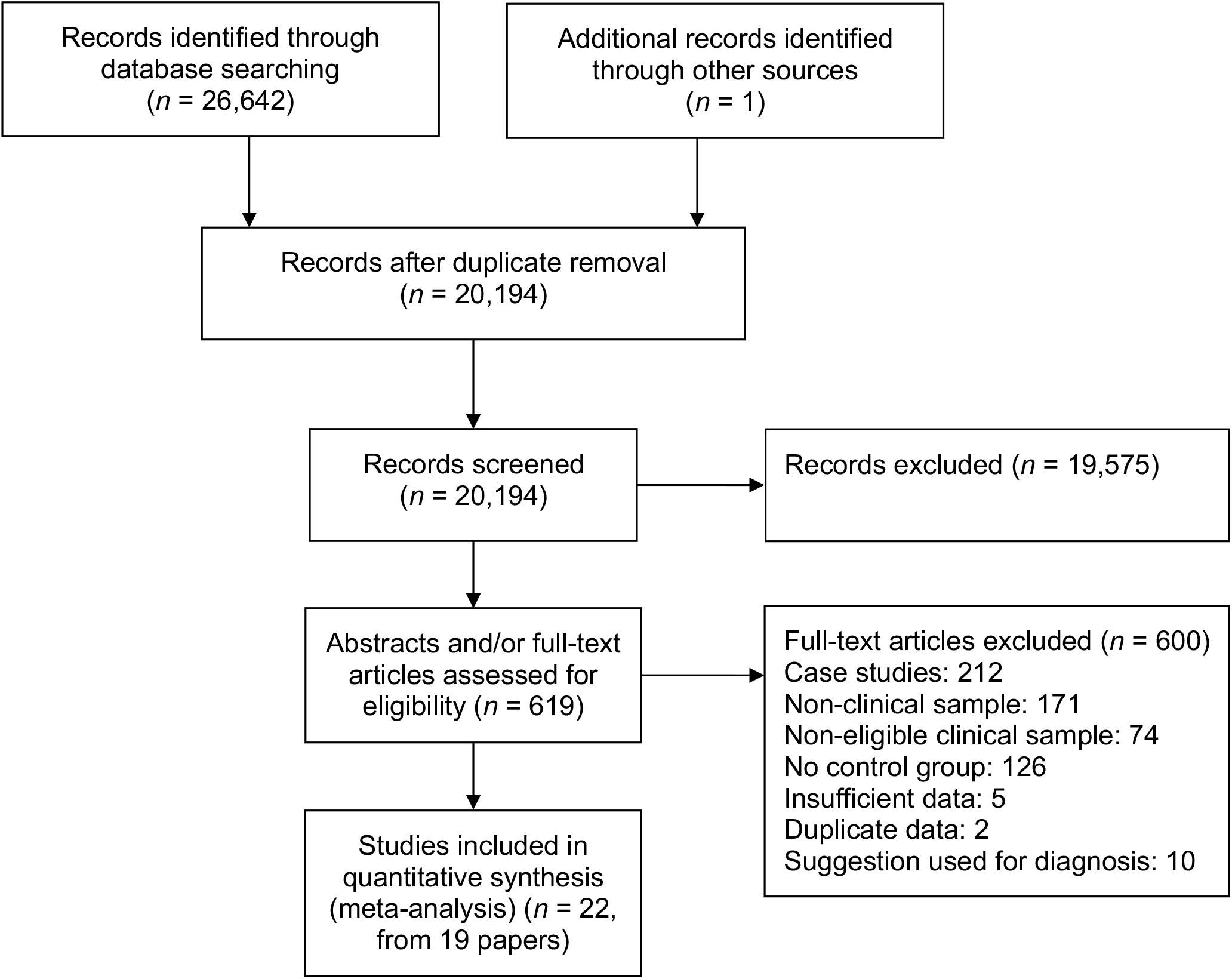
PRISMA flowchart of study selection process

**Table 1.**
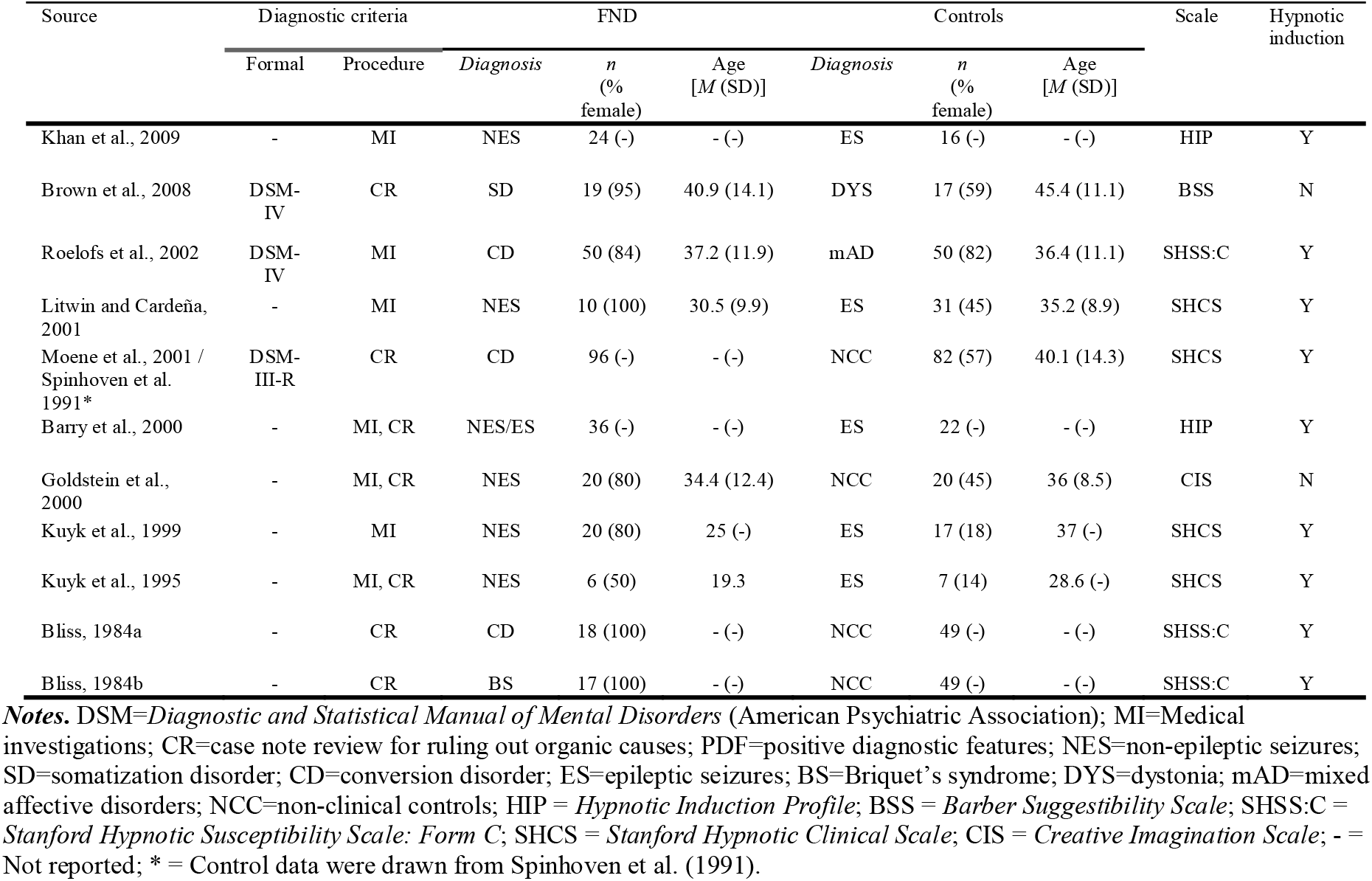
Characteristics of included studies measuring standardized suggestibility.

**Table 2.**
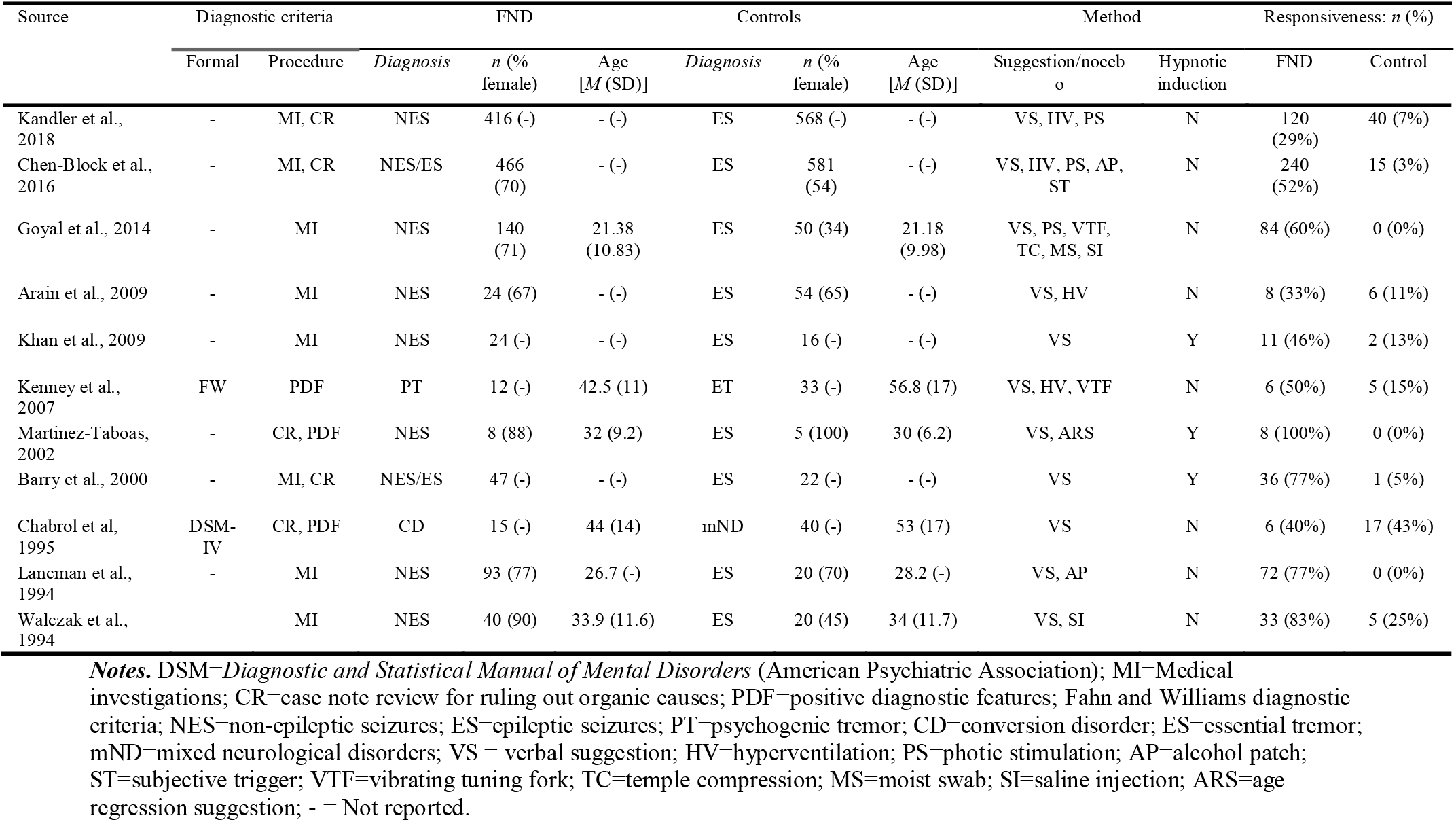
Characteristics of included studies measuring symptom suggestibility.

### Study and participant characteristics

The 19 papers included 11 standardized suggestibility studies (FND: *n* = 316, control: *n* = 360) and 11 symptom suggestibility studies (FND: *n* = 1285, control: *n* = 1409). The standardized studies were published between 1984 and 2009 and were conducted in the US (*k* = 5), the Netherlands (*k* = 4), and the UK (*k* = 2). The symptom studies were published between 1994 and 2018, and were conducted in the US (*k* = 7), Puerto Rico (*k* = 1), France (*k* = 1), India (*k* = 1) and the UK (*k* = 1). The mean percentage of females for standardized studies was 86.13 for FND (*k* = 8) and 45.71 for controls (*k* = 7), whereas for symptom studies (*k* = 6) it was 77.17 for FND and 61.33 for controls. In the standardized studies, the mean age for FND was 31.22 (*SD* = 8.01) (*k* = 6), and for controls it was 36.96 (*SD* = 5.09) (*k* = 7), whereas for the symptom studies (*k* = 6), mean age for FND was 33.41 (*SD* = 8.79), and 37.20 (*SD* = 14.38) for controls.

Details of the types of standardized scales and provocation methods as well as the use of a hypnotic induction are provided in **Table 1** and **Table 2**, respectively. A hypnotic induction was used in 9 standardized studies and 3 symptom studies. All symptom studies included verbal suggestion but varied in their use of various nocebo procedures.

### Methodological quality criteria

Ratings for each study on each of the 13 methodological quality criteria items are shown in **online supplemental content 1, Tables 1 and 2**. Although some of the criteria were met by the majority of studies, multiple criteria were not reliably met. Only 5 of 22 studies (23%) reported that the experimenter was blind to group, 18 (82%) described the inclusion/exclusion criteria, 16 (73%) described the diagnosis procedure and criteria in adequate detail, 10 (45%) described participant characteristics and 3 (14%) exhibited demographic comparability between patients and controls. In the standardized studies, 7 of 11 (64%) described the scale and procedure in adequate detail, and only 1 (9%) included a measure to correct for compliance. In the symptom studies, 8 of 11 (73%) clearly described the provocation method.

### Meta-analysis of standardized suggestibility

Meta-analysis of 11 standardized behavioural suggestibility studies found that patients with an FND exhibited greater suggestibility than controls, *SMD* = 0.48 [0.15, 0.81], *Z* = 2.84, *p* = .004 (see **Figure 2**). Positive results were observed in 8 studies with a high inconsistency of effect sizes across studies, *I*^2^ = 73%. Jackknife analysis in which each study effect was sequentially omitted and the analysis re-performed indicated that the group difference was reliably significant (*SMD* range: 0.40–0.56).

**Figure 2.**
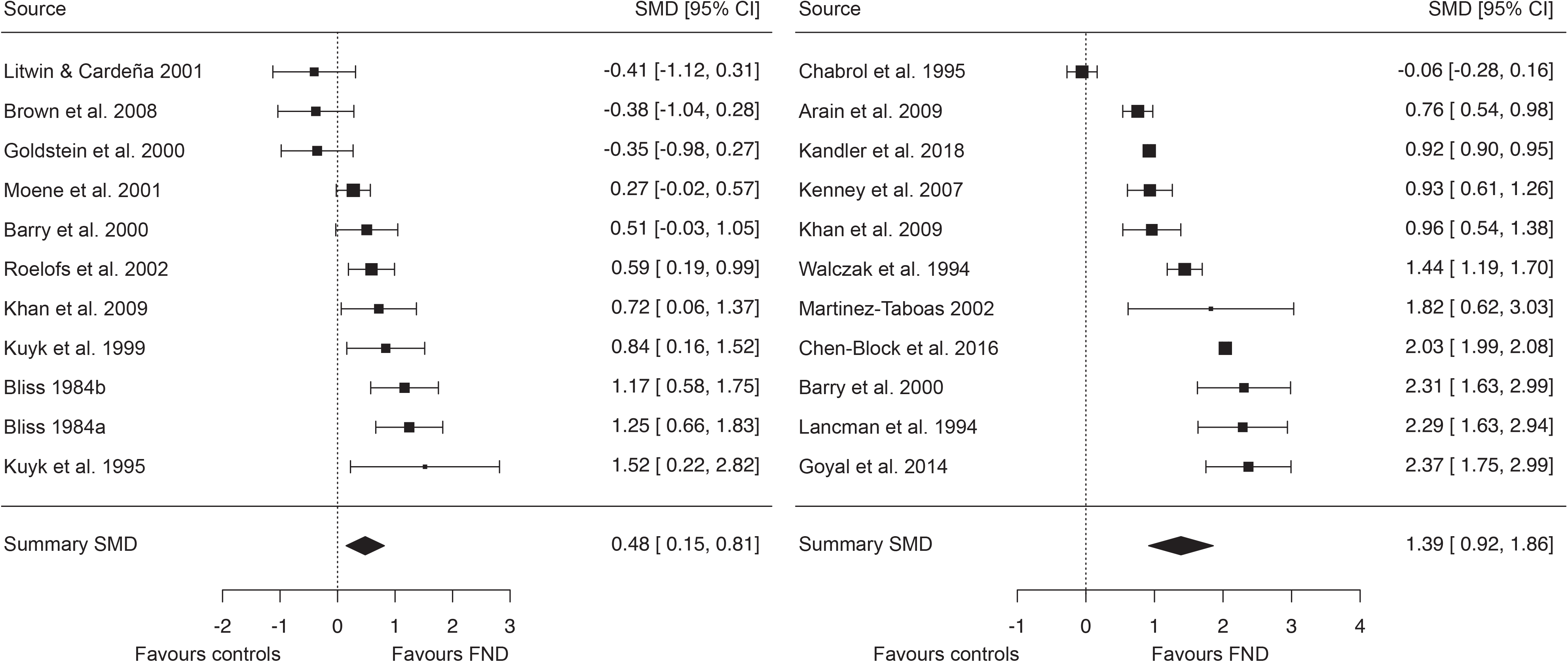
Forest plots of Standardised Mean Differences (*SMD*s) (with 95% confidence intervals) from (left) 11 standardized suggestibility studies and (right) 11 symptom suggestibility studies. Marker sizes reflect study weights with smaller markers denoting smaller weights.

### Meta-analysis of symptom suggestibility

Meta-analysis of 11 symptom suggestibility studies found that patients with FND displayed greater responsiveness than controls, *SMD* = 1.39 [0.92, 1.86], z = 5.77, *p*<.001 (see **Figure 2**). Overall positive responses were observed in 49% of patients with FND and 6% of controls (sensitivity = 49%, specificity = 94%). Ten of the 11 studies exhibited positive *SMD*s although there was substantial inconsistency in effect sizes across studies, *I*^2^ = 99%. Jackknife analysis revealed that the group difference remained significant after omitting each study in a sequential manner (*SMD* range: 1.25 to 1.54).

### Meta-analysis of standardized vs. symptom suggestibility

The weighted effect size for symptom studies was significantly greater than that for standardized studies, *z* = 2.60, *p* = .009. This difference remained stable, *z* = 2.29, *p* = .022, after removing the two studies included in both data sets. When the two data sets were aggregated, the cumulative standardised effect size was slightly less than 1, *SMD* = 0.96 [0.62, 1.29].

### Publication bias

Egger’s test did not suggest asymmetry in the distribution of effect sizes in the standardized studies, *z* = 0.69, *p* = .49, or in the symptom studies, *z* = 1.50, *p* = .13. A trim and fill estimate produced only a slight reduction in effect sizes for standardized studies, Δ*SMD* = –.05, and symptom studies, Δ*SMD* = –.10 (see **Figure 3**).

**Figure 3.**
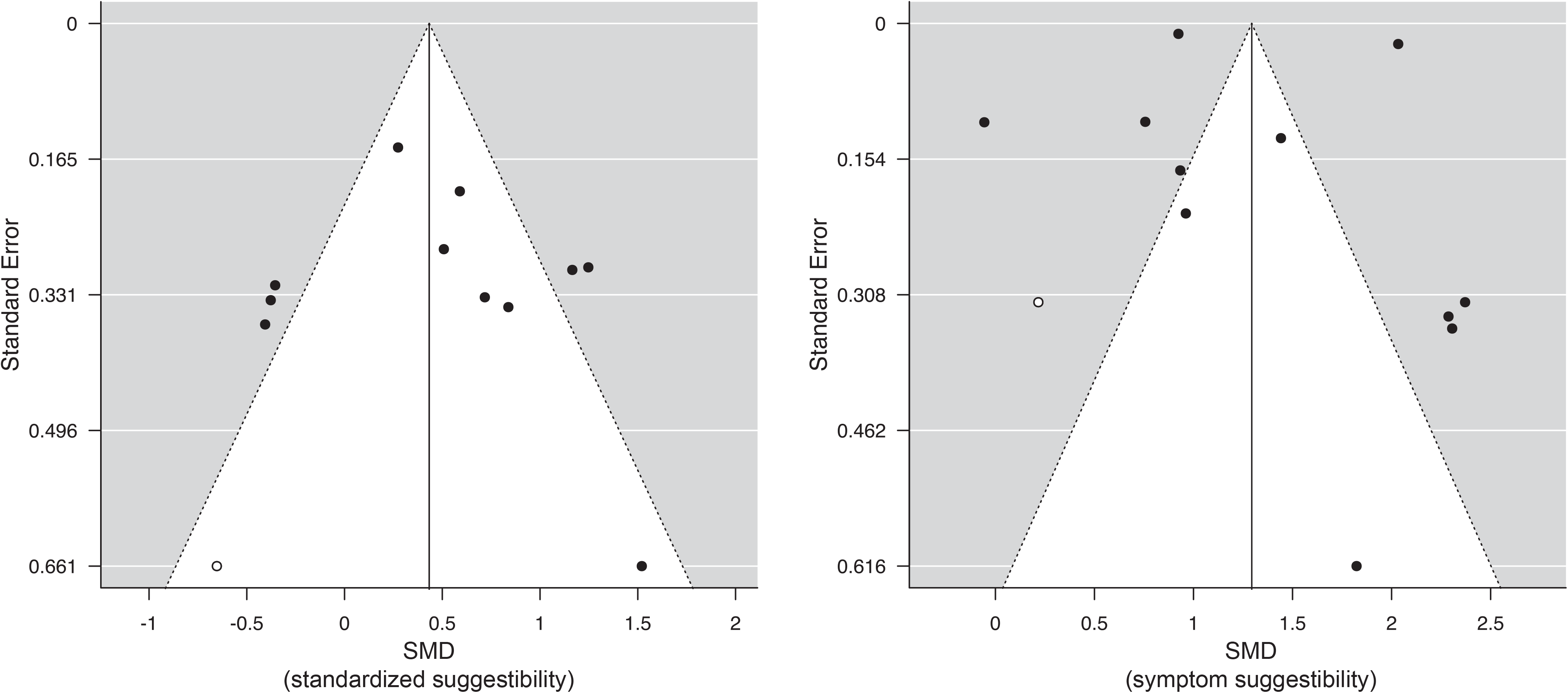
Funnel plots of standardized mean differences (*SMD*s) from (left) 11 standardised suggestibility studies and (right) 11 symptom suggestibility studies. Filled circles denote individual study effect sizes and empty circles denote estimated missing individual effect sizes due to publication bias imputed using the trim and fill method. Summary *SMD*s [95% CI] using the trim-and-fill method were *SMD* = 0.43 [0.10, 0.76] (standardized suggestibility studies) and *SMD* = 1.29 [0.84, 1.75] (symptom suggestibility studies).

### Meta-regression of standardized and symptom suggestibility

Given the observed inconsistency in the magnitude of effects, a set of meta-regression analyses considered whether group differences were moderated by binary and continuous predictors pertaining to study methodologies (see **Table 3**). Effect sizes were significantly larger when a hypnotic induction was included in standardized studies, but not in symptom studies. Effect sizes were also larger in studies that reported whether suggested symptoms were typical for the patient relative to those that did not report this information; this implies that effect sizes are not inflated by the inclusion of atypical symptoms in response rates for suggestive symptom induction. By contrast, effect sizes were not significantly related to the type of control (clinical vs. non-clinical; standardized studies) or suggestive induction protocol (suggestion vs. nocebo [suggestion and sham]; symptom studies). Experimenter blindness did not significantly moderate group differences with numerical differences in opposing directions for standardized and symptom studies. Similarly, methodological quality related to effect sizes in opposing directions: greater quality was significantly associated with lower effect sizes in standardized studies, but with larger (albeit non-significantly) effect sizes in symptom studies. These effects were primarily driven by procedure description. Standardized effect sizes were smaller in studies that met criteria for clarity of inclusion/exclusion criteria: *z* = –2.54, *p* = .011; diagnostic procedure: *z* = –2.82, *p* = .005; and scale administration procedure: *z* = –2.28, *p* = .022 (all other *p*s>.09). In contrast, symptom effect sizes were larger in studies that met criteria for clarity of diagnostic procedure, *z* = 3.23, *p* = .001 (all other *p*s>.09).

**Table 3.** Meta-regression analyses for standardized and symptom suggestibility studies (*SMD* [95% CIs] (*k*))

| Moderator |  | Analysis groups |  | *Z* | *p* | *I*<sup>2</sup> |
| --- | --- | --- | --- | --- | --- | --- |
| Suggestibility type |  |  |  |  |  |  |
| **Hypnotic induction** |  | No | Yes |  |  |  |
|  | Standardized | -0.37 [-0.82, 0.09] (2) | 0.66 [0.34, 0.97] (9) | 2.74 | .006 | 61% |
|  | Symptom | 1.30 [0.76, 1.85] (8) | 1.66 [0.67, 2.64] (3) | 0.62 | .54 | 100% |
| **Experimenter blindness** |  | Unblind | Blind |  |  |  |
|  | Standardized | 0.46 [0.03, 0.89] (7) | 0.54 [-0.09, 1.16] (4) | 0.20 | .84 | 76% |
|  | Symptom | 1.44 [0.94, 1.93] (10) | 0.93 [0.61, 1.26] (1) |  |  |  |
| **Suggestive symptom induction method** |  | Suggestion | Nocebo |  |  |  |
|  | Symptom | 1.20 [0.11, 2.28] (4) | 1.50 [0.93, 2.08] (7) | 0.69 | .49 | 100% |
| **Suggestive symptom typicality** |  | Not reported | Typical |  |  |  |
|  | Symptom | 0.60 [-0.14, 1.34] (3) | 1.70 [1.14, 2.26] (8) | 2.07 | .038 | 100% |
| **Control sample type** |  | Non-clinical | Clinical |  |  |  |
|  | Standardized | 0.58 [-0.08, 1.24] (4) | 0.42 [0.01, 0.82] (7) | -0.40 | .69 | 76% |
| **Methodological quality** |  |  |  |  |  |  |
|  | Standardized | 12-item scale |  | -2.21 | .027 | 66% |
|  | Symptom | 10-item scale |  | 1.76 | .079 | 99% |

## Discussion

The results of this meta-analysis suggest that FND patients display elevated suggestibility on standardized behavioural scales and in response to suggestive symptom induction protocols, consistent with theoretical predictions to this effect.^7, 20, 21^ There was no evidence for publication bias although there was considerable heterogeneity in both data sets, which was partly explained by methodological variability.

These findings are consistent with models proposing responsiveness to suggestion as a vulnerability factor for FND and those attributing functional symptoms to precise symptom priors.^8, 13^ Recent theoretical work conceptualizes functional symptoms as arising from an interaction between multiple factors including an autonomic arousal response, inhibitory deficits that increase the likelihood of body misrepresentation, and symptom priors that collectively trigger the automatic activation of rogue symptom representations or conditioned actions.^8^ Suggestibility may confer heightened sensitivity to symptom-specific cues or dissociative responses to stressors. Moreover, suggestibility has been proposed to reflect a generalized tendency to form precise priors that override various motor and perceptual systems,^31^ which is thought to be a key process in FND^13^ and symptom reporting more generally.^14^ This aligns with research showing that hypnotic suggestibility predicts symptom severity in FND patients.^32^ Observed links between hypnotic suggestibility and emotional responsiveness to social cues,^23^ highlight the potential role of suggestibility in symptom modeling or triggering through social observation.^8^ The perception that functional symptoms are extra-volitional may be further augmented by aberrant meta-awareness of intentions, as observed in both FND^11, 12^ and high hypnotic suggestibility.^9, 10^

Although the two forms of suggestibility moderately covary,^33^ FND patients were more responsive to symptom-specific (*SMD* = 1.39 [0.92, 1.86]) than to standardized suggestions (*SMD* = 0.48 [0.15, 0.81]), implying selectively greater suggestibility for functional symptoms. Indeed, the sole non-significant provocation study^34^ administered suggestions for generic symptoms that did not necessarily mirror patients’ symptom profiles. Previous research similarly found that highly dissociative individuals and dissociative and acute stress disorder patients are more responsive to suggestions for dissociative experiences (e.g., amnesia).^19^ Symptom suggestibility in FND may thus partly reflect a kindling process whereby symptoms become more responsive to verbal suggestion over time.^8^

Atypical suggestibility in FND patients complements research showing greater suggestibility in germane conditions, such as dissociative and stress disorders^19, 20^ and somatoform disorders.^35^ Many of these conditions are characterized by detachment and/or compartmentalization symptoms.^36^ Variability in such symptoms may account for heterogeneity in the suggestibility profiles of FND patients, with heightened responsiveness to suggestions being specific to, or more pronounced among, patients with marked dissociative symptomatology.^19, 37^ Moderation analyses indicated that the administration of a hypnotic induction was associated with greater standardized suggestibility among FND patients. This is consistent with the proposal that individuals with compartmentalization symptoms benefit more from a hypnotic induction^19^ although the mechanistic basis of this effect remains unclear.^23^

These effects attest to the efficacy of suggestion in the diagnosis of FND.^15^ Suggestive symptom induction displayed high specificity (94%) although sensitivity is poor (49%), indicating that this technique is insufficient as a standalone diagnostic procedure. The inclusion of sham methods or a hypnotic induction were not associated with greater discrimination of FND patients and controls relative to verbal suggestion alone but warrant further attention. Suggestive symptom induction is likely to be especially valuable in suggestible populations such as adolescent and elderly samples^23^ or patients with comorbid dissociative or stress disorder diagnoses.^19, 20^ It may also inform diagnosis of comorbid non-epileptic seizures in epilepsy patients^38^ and medication prescription in FND patients.^39^ Insofar as suggestibility is a positive predictor of outcome with suggestion-based treatments,^18^ these results also support greater incorporation of suggestion techniques in treatment protocols, which show promising results in randomized-controlled trials.^16^ However, they also highlight the need to control for suggestion, or consider its role, in diagnostic and treatment procedures, particularly those that evoke strong response expectancies.

## Limitations

The principal limitations of these data concern methodological variability across studies. Methodological quality was significantly or descriptively related to effect sizes in both data sets albeit in opposite directions. Among standardized studies, older studies that did not precisely specify inclusion/exclusion criteria and/or diagnostic and scale administration procedures tended to exhibit larger effect sizes; in contrast, precise specification of diagnostic procedures was associated with larger effect sizes in symptom studies. In most studies, the operator administering the assessment was not blind to patient group, which may inflate effect sizes^22^, although there was no evidence for an experimenter effect in the standardized studies. The sole blind symptom study had a lower effect size than the remainder of the studies but was still large in magnitude (*SMD* = 0.93 [0.61, 1.26]). Most studies included clinical controls (e.g., epilepsy patients), which raises issues regarding generalizability although effect sizes did not significantly relate to control type. Symptom suggestibility estimates are also confounded by baseline symptom frequency, which is not incorporated into these assessments.^15^ This potentially renders patients with high symptom frequency at an increased risk of false positive responses, although there is evidence that this is not the case.^40^ The studies also varied in whether successful responses to symptom induction protocols were contingent upon the typicality of the response, which accounted for variability in effect sizes. Standardized studies were limited insofar as only one controlled for compliance. In addition, standardized suggestibility scales include a disproportionate number of suggestions for dissociative and functional symptoms (e.g., paralysis),^20, 23, 32^ raising the question of whether elevated suggestibility in FND generalizes beyond these symptoms. Collectively, these findings underscore the need for optimization and standardization of suggestive symptom induction protocols,^15^ compliance-correction,^23^ and more diverse suggestion batteries.

## Conclusions

This meta-analysis corroborates the long-held view that FND is characterized by elevated suggestibility. Increased suggestibility has direct implications for the risk factors underlying this condition, the use of suggestion to aid diagnosis, the utility of suggestion-based treatments for functional symptoms, and heterogeneity within this population.

## Data Availability

This study reports a meta-analysis. All data are available in the paper, the supplementary materials, or the original papers.

## Acknowledgements

The authors thank Che Ofuasia, Goldsmiths, University of London, for contributing to the data coding and evaluation.

## Contributors

All authors conceived the project. LW carried out the database searches and data coding with assistance from RJB, TT, and DBT. LW and DBT performed the meta-analysis with assistance from RJB and TT. LW and DBT drafted the initial manuscript. All authors reviewed and approved the final version of the manuscript.

## Patient consent for publication

Not required.

## Provenance and peer review

Not commissioned; externally peer reviewed.

